# Dynamic Prioritization of COVID-19 Vaccines When Social Distancing is Limited for Essential Workers

**DOI:** 10.1101/2020.09.22.20199174

**Authors:** Jack H. Buckner, Gerardo Chowell, Michael R. Springborn

## Abstract

COVID-19 vaccines have been authorized in multiple countries and more are under rapid development. Careful design of a vaccine prioritization strategy across socio-demographic groups is a crucial public policy challenge given that (1) vaccine supply will be constrained for the first several months of the vaccination campaign, (2) there are stark differences in transmission and severity of impacts from SARS-CoV-2 across groups, and (3) SARS-CoV-2 differs markedly from previous pandemic viruses. We assess the optimal allocation of a limited vaccine supply in the U.S. across groups differentiated by age and also essential worker status, which constrains opportunities for social distancing. We model transmission dynamics using a compartmental model parameterized to capture current understanding of the epidemiological characteristics of COVID-19, including key sources of group heterogeneity (susceptibility, severity, and contact rates). We investigate three alternative policy objectives (minimizing infections, years of life lost, or deaths) and model a dynamic strategy that evolves with the population epidemiological status. We find that this temporal flexibility contributes substantially to public health goals. Older essential workers are typically targeted first. However, depending on the objective, younger essential workers are prioritized to control spread or seniors to directly control mortality. When the objective is minimizing deaths, relative to an untargeted approach, prioritization averts deaths on a range between 20,000 (when non-pharmaceutical interventions are strong) and 300,000 (when these interventions are weak). We illustrate how optimal prioritization is sensitive to several factors, most notably vaccine effectiveness and supply, rate of transmission, and the magnitude of initial infections.

## 1 Introduction

As the novel coronavirus (SARS-CoV-2) continues to inflict substantial morbidity and mortality around the world despite intervention efforts, public health experts see a vaccine as essential to dramatically reduce the mortality burden and possibly halt local transmission (1). Novel coronavirus disease 2019 (COVID-19) has resulted in over 1.5 million confirmed deaths globally (2) as of mid-December 2020. Fortunately, multiple promising vaccines are under rapid development, with the final weeks of 2020 seeing the first authorization and shipping of doses (3). However, vaccine availability will be highly constrained for at least several months (4). This scarcity, combined with stark differences in the spread and impact of SARS-CoV-2 across demographic groups, means that prioritization of the vaccine poses a public health challenge, and as such under active discussion by the “Advisory Committee on Immunization Practices (ACIP) of the US Centers for Disease Control and Prevention (CDC) and the National Academy of Medicine (NAM), as well as globally at the World Health Organization (WHO) and elsewhere” (5).

An effective public health policy for pandemic vaccine allocation requires an understanding of how risk of infection and severe disease varies across socio-demographic groups and how a given vaccine policy will impact the continued spread of infections within the population. Accounting for these two processes is critical when the population with the greatest risk of infection differ from those with the greatest risk of severe disease, as is the case for COVID-19, because an effective policy will need to balance direct protection of the most vulnerable against limiting secondary infections and rapidly achieving herd immunity (6). These key components can be integrated into a mathematical and statistical modeling framework of the transmission dynamics of the novel pathogen. Such an analytic framework can then be utilized to investigate the optimal vaccine allocation strategies to achieve a defined public health objective while taking into account the value of vaccines for mitigating health outcomes at the individual and population level. Previous experience with vaccine development mid-pandemic offers limited insights for SARS-CoV-2 prioritization.

SARS and Zika vaccine development was incomplete when those outbreaks ended (7). In 2009, as the novel A/H1N1 influenza virus continued to spread across the U.S., researchers investigated optimal vaccination strategies using an age-structured dynamical model. They found that school-aged children and their parents should be prioritized, a strategy that would indirectly protect individuals at higher risk of severe health outcomes (8). Sharp differences in the epidemiology of human influenza and COVID-19 indicate that vaccination strategies against the ongoing pandemic should not simply mirror vaccination policies against influenza. For example, COVID-19 is associated with lower susceptibility to infection among children and adolescents (9, 10) and has a substantially higher infection fatality rate overall that also increases markedly with age (11). Toner et al. (5, p. 24) provide a detailed overview of the 2018 pandemic influenza vaccination plan and conclude that, “the priority scheme envisioned…does not comport with the realities of the COVID-19 pandemic and new guidance is needed.”

We develop and apply a mathematical model to assess the optimal allocation of limited COVID-19 vaccine supply in the U.S. across socio-demographic groups differentiated by age and essential worker status (see Methods). The transmission dynamics are modeled using a compartmental model tracking eight demographic groups through the nine disease states as shown in Fig. 1. The parameters are calibrated to capture our current understanding of the epidemiology of COVID-19, and our analysis is designed to capture two key features of COVID-19 prioritization: essential workers and the gradual availability of vaccines over time. A large number of workers are constrained in their ability to work from home (essential workers) exposing them to higher level of risk of infection, and increasing the chance they transmit the disease if infected. Policies that account for the greater risk essential workers are exposed to may be more just and highlight a group of individuals “who have been overlooked in previous allocation schemes” (5). Furthermore, these policies may be more effective at mitigating morbidity and mortality as they can account for a key factor driving transmission of the disease.

**Figure 1:**
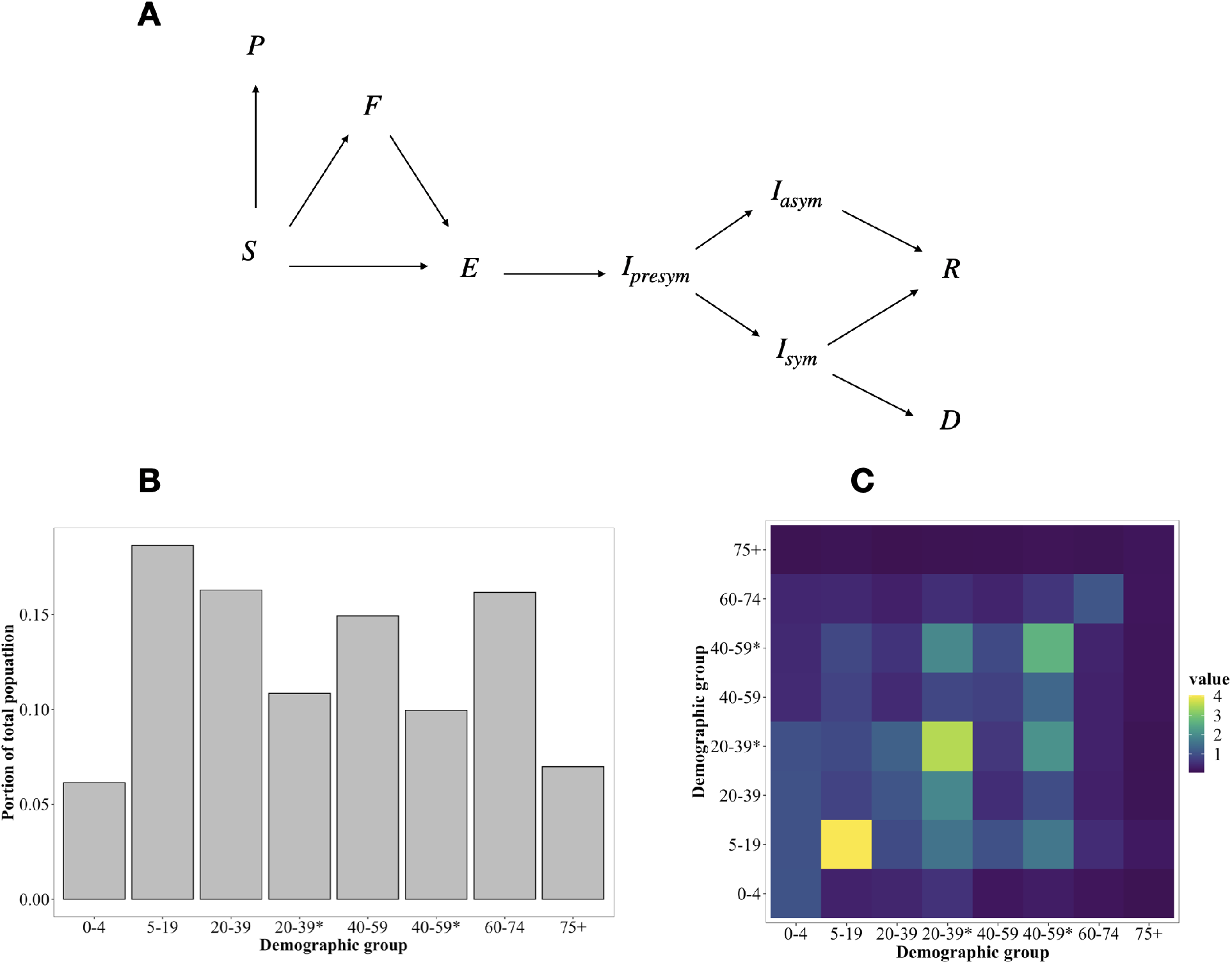
Schematic of the modeled movement of individuals between epidemiological states (A), the portion of individuals from the U.S. population in each demographic group determined by essential worker status (*) and age (B), and the contact rates between demographic groups, given by average daily number of contacts a group on the horizontal axis makes with a group on the vertical axis (C).

To account for the gradual roll out of vaccines we employ stochastic non-linear programming techniques to solve for vaccine prioritization policies that distribute vaccine to susceptible individuals and change on a monthly time step responding to changes in the epidemiological status of the population (shares of the population in different disease states). These dynamic policies account for a key feature of the policy making process since the supply of vaccine is likely to be constrained with available doses administered as they become available over a period of several months.

The transmission of COVID-19 is a complex process contingent on the characteristics of the disease and ever changing social behavior. Furthermore many of the key dynamics can change depending on the spatial scale considered, with differences in the transmission process within and between communities. We seek to summarize the features of the complex and evolving processes that are most relevant to the spread of the disease within and between socio-demographic groups. To do so we model COVID-19 transmission with the social contact hypothesis (12) and describe the contact patterns between demographic groups using contact matrices estimated for the United States from Prem et al. (13) scaled by the location where the contacts were made (home, school, work and other) to reflect the impacts of social distancing. Although these assumptions present a stylized version of contacts during the pandemic, they allow us to capture many key feature of social contacts, such as the concentration of contacts within age groups, parent-child relationships and receiver-caregiver relationships (14).

To our knowledge there are no published analyses of optimal COVID-19 vaccination prioritization. Notable analyses in preprint form include Matrajt et al. (15), Bubar et al. (16) and Hogan et al. (17).^1^ All consider the optimal allocation of vaccines across five or more age groups within a country. Their approaches feature rich exploration of policy sensitivity to vaccine effectiveness and availability. Matrajt et al. is particularly detailed in this respect, while Bubar et al. uniquely consider differences in demographics and contact rates across multiple countries and Hogan et al. also consider allocation between countries. Our analysis is differentiated by a deeper approach to the behavioral, demographic and decision models by addressing social distancing, essential worker groups, and allocation policies that can change over the course of the vaccination campaign.

General ethical guiding frameworks for vaccine prioritization decision-making have appeared earlier in the literature. Toner et al. (5) emphasize promoting three ethical values: the common good; fairness and equity; and legitimacy, trust and communal contributions to decision-making. Emanuel et al. (4) promote four ethical values: maximizing benefits, treating equally, instrumental value, and priority to the worst off. Our analytic focus on minimizing new infections, years of life lost (YLL), or deaths emerges from promoting “the common good” or “maximizing benefits”. Our focus on essential worker groups illustrates how ethical values (e.g. prioritizing essential workers due to the fairness of protecting those placing themselves at risk) may overlap with the common good (e.g. prioritizing essential workers to best reduce mortality and transmission). Issues of fairness and equity and protecting the worst off are not directly analyzed here but remain critical considerations.

For the sake of simplicity, we do not address in detail the potential set of complex and differential feedback processes between health status and opening of schools, workplaces and other institutions. While we limit policy objectives to a concise metric of health outcomes (minimizing expected cases, YLL, or deaths) we acknowledge that other values of returning to school, work and social life are important. Finally, we do not address additional vaccine complications, such as temporary effectiveness, potential side effects or any failure to take a second dose of the vaccine if necessary.

Although much is known about the epidemiology of COVID-19, uncertainty remains a key limitation to modeling the disease. Therefore, we consider a wide range of plausible scenarios and focus on the general features of the solutions, the commonalities between the alternative scenarios, and identification of model parameters drive systematic differences in optimal vaccine allocations.

Given these assumptions we find that optimal allocation strategies are responsive to both the initial and evolving epidemiological landscape of the disease. When the focus is minimizing deaths or YLL, we find that optimal allocations target essential workers and seniors (ages 60+). Alternatively, When infections are minimized, essential workers are prioritized followed by school age children across a range of likely scenarios. We find that prioritization can substantially improve public health outcomes—31 to 40% in the Base scenario, relative to untargeted vaccination. Two components unique to our model are important contributors to this improvement. First, policies that differentiate and target essential workers in addition to age substantially outperform those utilizing age-alone. Furthermore essential worker differentiation reduces trade offs between objectives (e.g. deterioration of YLL and infection metrics when focused on minimizing deaths). Second, extending from a static allocation (without phases) to allowing changes in prioritization over time provides substantial gains. Finally, while optimal prioritization is quite insensitive to model specification when minimizing infections, we find some sensitivity when focused on minimizing deaths or YLL. This sensitivity indicates benefits to adjusting the targeting strategy at the local level to match epidemiological conditions.

## 2 Results

We first present results from a single “Base” scenario, representing a plausible set of parameters, to illustrate the qualitative nature of optimal dynamic prioritization. These results are then compared to a set of alternative model scenarios as described in Table 1. In Fig. 2 the Base model allocation decisions are shown for each monthly decision period (in percent of vaccine supply) and then cumulatively (in percent of group vaccinated) at three and six months, respectively. Broadly, we find that the optimal policy is very dynamic: specific groups are targeted each period and these targets shift over time. Furthermore, targeting is very narrow initially but then becomes less so as a larger fraction of the population has been covered.

**Table 1:** Descriptions of alternative scenarios to the Base model (see SI Appendix A.3 for specific levels).

| Scenario | Change from Base scenario parameters | Source |
| --- | --- | --- |
| Base scenario | None (Base parameter values are provided in SI Appendix A.2) |  |
| High initial infections | Increased number of initial symptomatic infections (300% increase) | Assumed: pandemic state will vary between localities when vaccine first available to the general public. |
| Strong NPI | Non-social distancing (NSD) NPI are strong, resulting in a declining infection rate | Consistent with $R < 1$ |
| Weak NPI | NSD NPI are weak, resulting in a sharply increasing burden of infection | Consistent with $R \gg 1$ |
| Weak vaccine | Lower vaccine effectiveness (success rate) for all age groups relative to the Base scenario | Minimum value required by FDA guidelines |
| Weak vaccine 60+ | Lower vaccine effectiveness for ages 60+ yrs. | Informed by influenza vaccine effectiveness |
| Even susceptibility | All ages are equally susceptible to infection. Increase in susceptibility for ages $< 20$ yrs relative to Base | Assumed: tests sensitivity to age-dependent susceptibility described by (9) and (19) |
| Low supply | Sufficient supply for 5% of the population monthly (50% of supply relative to Base scenario; prioritization changes every 10% of the population vaccinated, such that decision period is 2 months) | Assumed: vaccine supply is uncertain and known to impact optimal allocations (20) |
| Ramp up | Vaccine supply is 5% per month for the first two months and 10% per month thereafter (first decision period is 2 months so increments of 10% of the population are vaccinated each decision period) | Informed by comments from the scientific head of the U.S. vaccine development program (21) |
| Open schools | Rate of social contact in schools increased from 30% in Base model to 70% | Assumed: tests sensitivity of optimal allocations to school closure intensity |
| High contacts | Increased number of contacts outside the home, school and workplace (50% increase relative to base) | Assumed: tests sensitivity to relaxed distancing |

**Figure 2:**
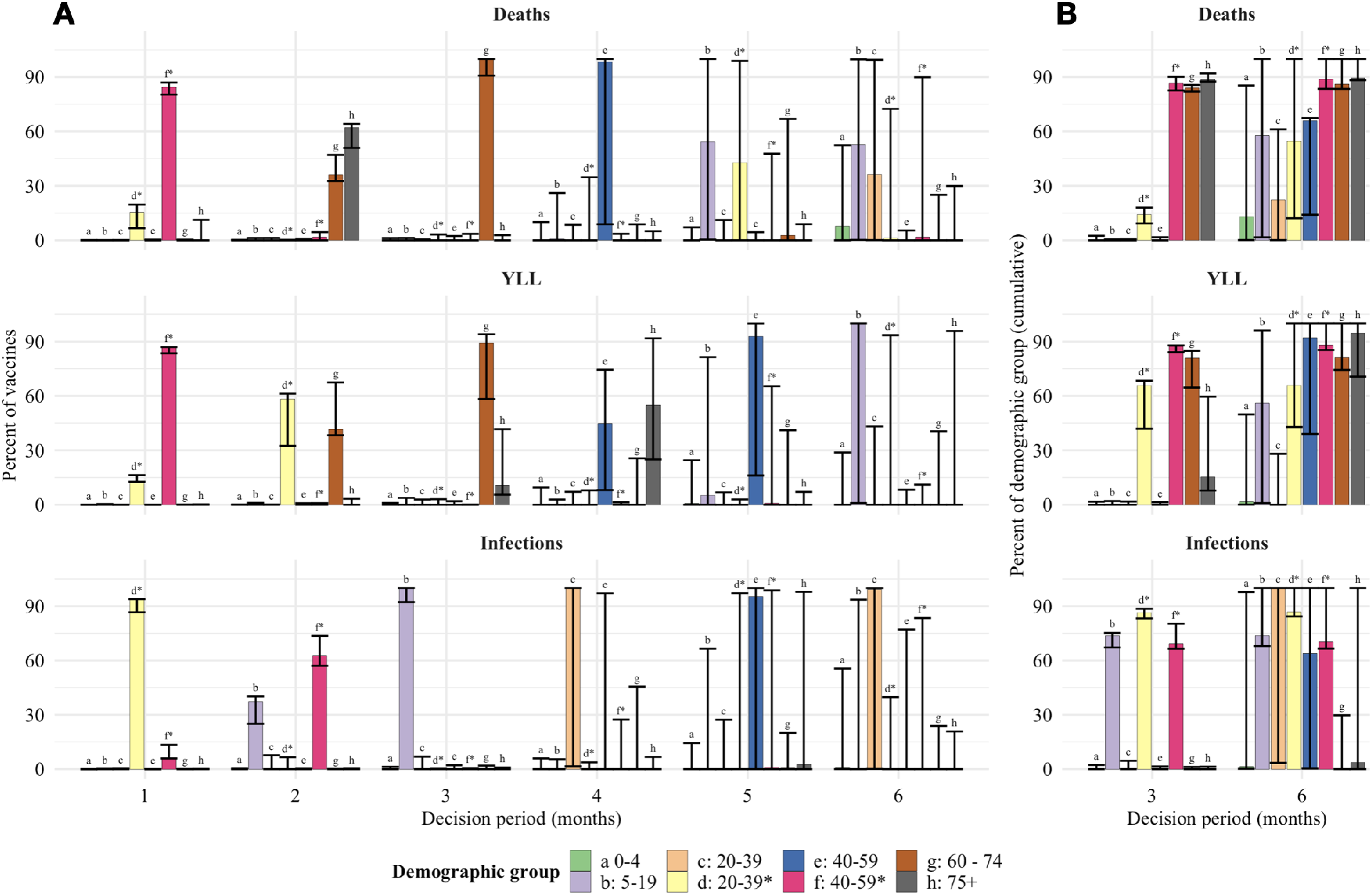
The optimal allocation of vaccines (vertical axes) between demographic groups for each decision period (horizontal axis) under the Base scenario (A). The three rows represent each objective, to minimize deaths, minimize years of life lost (YLL) and minimize infections. The bars for the six decision periods show the percentage of vaccines allocated to a specific group (indicated by a letter, color, and stars indicating essential worker groups) in that period. The two final columns (B) show cumulative measures at the end of months three and six, respectively, for the percent of each group that has been vaccinated. The whiskers on each bar represent the sensitivity of the optimal solution to small deviations in the outcome, specifically the range of allocations resulting in outcomes within 0.5% of the optimal solution.

The whiskers on bars in Fig. 2 show the range of alternative allocations that still produce an outcome that is within 0.5% of the optimum. These indicate that the optimized outcome is relatively sensitive to substitutions between groups for the first three months as indicated by narrow whiskers around the cumulative allocations. There is, however, some limited ability to substitute vaccines between the two essential worker groups in the first two months when minimizing YLL or deaths. As the size of the susceptible population declines due to vaccination and infections the optimized outcomes become less sensitive to substitutions (longer whiskers) with shifts between nearly all groups possible without substantial sacrifice. This suggests that targeting strategies can become less strict over time as the most vulnerable populations are protected. Comparing individual periods (Fig. 2A) and cumulative measures (Fig. 2B) shows that whiskers represent a combination of substitution between groups as well as between periods for the same group.

Across objectives there are substantial differences in which groups are targeted early on. When minimizing deaths, targeting progresses from essential workers (20-39*, 40-59*), to the oldest (75+) and then younger seniors (60-74). These groups are a mix of those at high risk of mortality (older groups) and high risk of contraction and spread (essential workers). When minimizing YLL, younger seniors are targeted earlier (given their longer average years of life remaining). Finally, when minimizing infections we find that younger essential workers take top priority, followed by older essential workers and school-age children (5-19), since these groups have higher contacts and thus risk of contraction and spread.

In Fig. 3A we show the dynamic path of infections, starting from the period in which vaccines become available, under various policies. As expected, infections are highest given no vaccines. Results for allocating vaccines in a manner proportional to each group’s size shows the substantial value of even “untargeted” vaccines. As expected, the policy for minimizing infections leads to the lowest level of infections.

**Figure 3:**
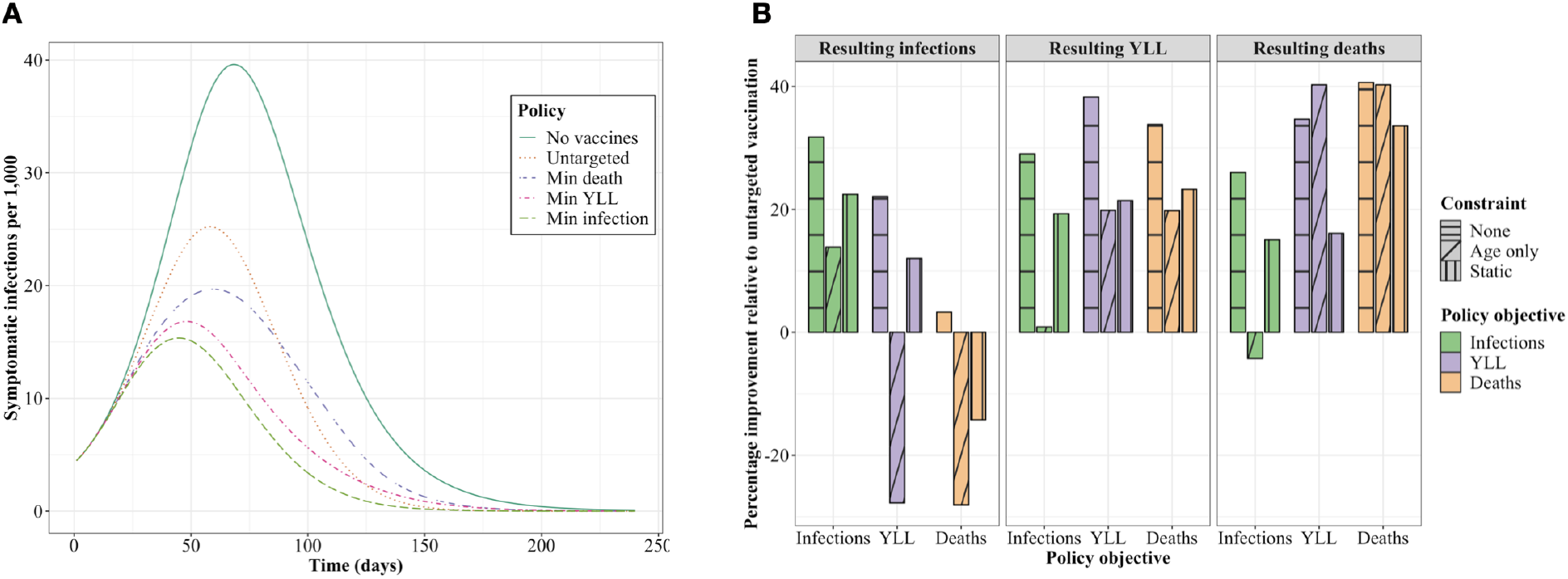
The number of infections per 1,000 individuals over time under reference policies (no vaccines; untargeted vaccine allocation) and optimized policies minimizing a given metric (A); and the performance of each optimized policy relative to an untargeted allocation policy (B) for the Base scenario. The bars are boxed by each resulting metric, colored by the objective driving each policy and textured to reflect any constraint considered (e.g. age-only or static policies).

In Fig. 3B we show the performance of various policies for resulting outcome metrics (infections, YLL and deaths) in terms of the percentage improvement relative to an untargeted vaccine allocation. We consider the optimal policies presented in Fig. 2 where the objective is minimizing infections (green), YLL (purple), or deaths (orange) with no constraints (“none”). We also consider two constrained alternatives: an “age-only” dynamic policy that does not differentiate by essential worker status, and a “static” policy where the fractional allocation across groups does not change over decision periods.^2^ We find that the unconstrained policy—that is dynamic and differentiated by essential workers—outperforms the untargeted approach by approximately 31-40% depending on the objective. Relative to the unconstrained policy, the age-only and static polices perform substantially worse for infections and YLL, though not for deaths. However, even while the age-only and static polices do not substantially impede performance in minimizing deaths, these constrained approaches still suffer substantial performance loss (9-18 percentage points) in the other two outcomes not optimized (YLL and infections) but clearly still of interest.^3^ In other words, accounting both for essential workers and a dynamic prioritization strategy provides substantial improvements in the metric being optimized and/or the other two metrics of interest.

In general, we find that there are trade offs between the objectives. For example policies that minimize infections result in significantly more deaths than a policy that minimizes deaths. However, we find that differentiating essential workers substantially reduces these tradeoffs between objectives relative to age-only or static polices.

### 2.1 Sensitivity of vaccine prioritization

To assess how robust our Base scenario findings are to key uncertainties in the model, we conduct three different sensitivity analyses. First we consider a set of 10 alternative plausible scenarios involving a broad set of model inputs; then we focus on a narrower set of 4 parameters each explored in richer gradient detail; finally we examine a few fundamental changes to model structures.

#### 2.1.1 A broad set of alternative scenarios

We solved for the optimal vaccine allocation across a range of 10 alternative scenarios selected to assess sensitivity to key assumptions of the Base model. Differences between these scenarios and the Base case are detailed in Table 1. Relative to the Base model, in these alternative scenarios we consider: higher initial infections; stronger or weaker non-social distancing non-pharmaceutical interventions (NPI) like mask wearing; weaker vaccine effectiveness overall or for seniors (60+); lower vaccine supply or supply that starts low and ramps up; more open schools; or higher contact rates overall.

To compare and contrast optimal early vaccination allocation for each scenario and objective, in Fig. 4A we show the percentage of each group vaccinated after 30% of the overall population is covered (typically in three months, except for alternative supply scenarios). We find that high priority groups—by percent of group vaccinated—are typically but not always robust to the alternative scenarios. For example, when deaths are considered (Fig. 4A, top panel) we see substitution between younger essential workers (20-39*) and ages 60-74 and when YLL are considered there is substitution between younger essential workers and ages 75+.

**Figure 4:**
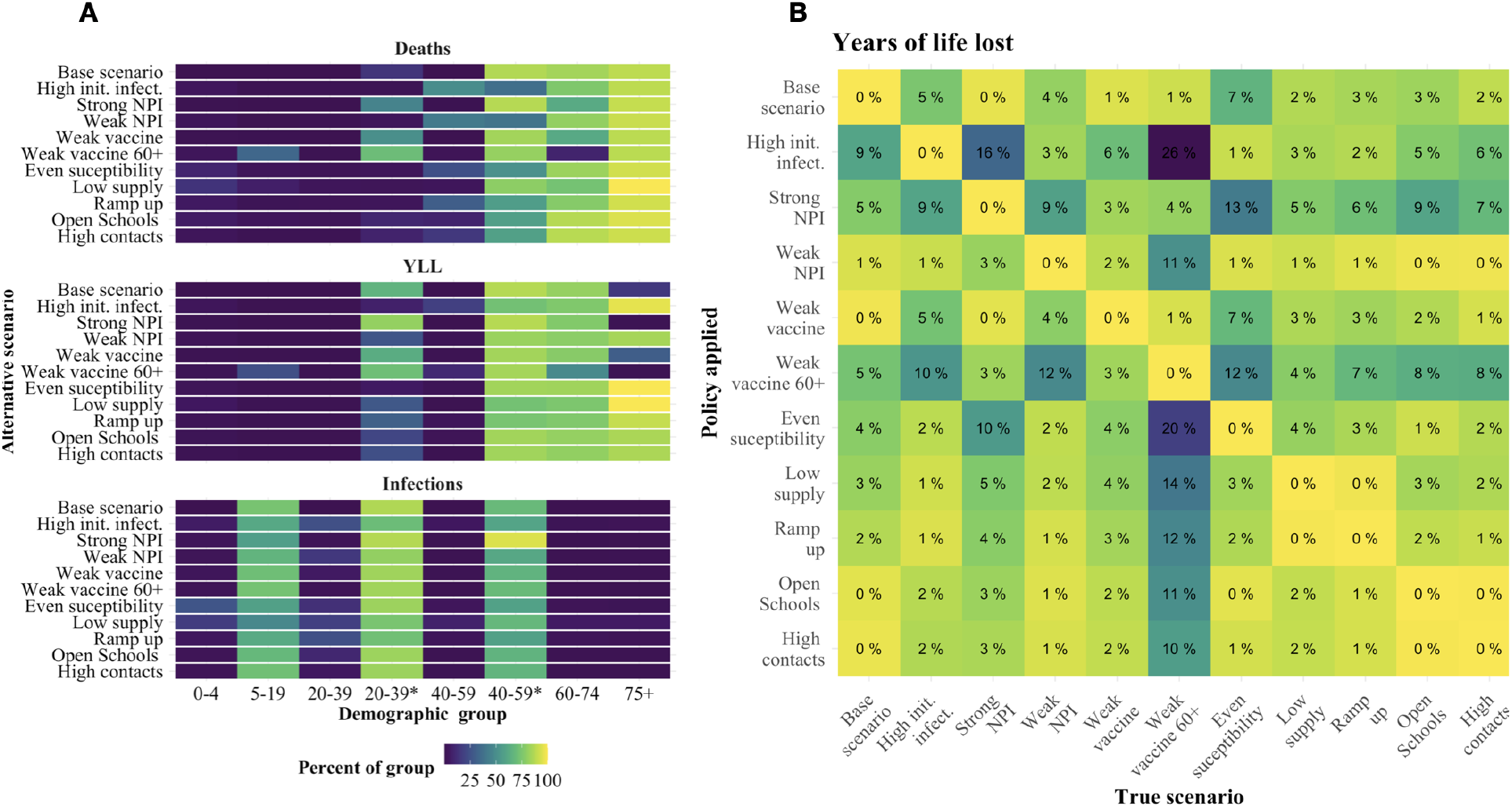
The cumulative percent of each demographic group (horizontal axis) vaccinated after the first 30% of the population is vaccinated under the alternative scenarios (vertical axis) and each objective (panel) (A). The percentage of additional YLL in excess of the optimum when applying a policy for a given alternative scenario (row) when a particular scenario is the “truth” (column) (B).

For insight into the cost of error in specifying the correct scenario, we assessed the performance of the policy identified for each of the 11 alternative scenarios, depending on which of these 11 is the “true” scenario. In Fig. 4B we show these results for the YLL objective. For example, the first column shows the performance loss (in percentage of additional YLL above the optimum) when the true scenario is the Base model but the decision maker applies a policy matched to any of the alternative scenarios (rows). By construction, when the policy applied matches the true scenario, the performance loss is zero. When YLL is the focus and the Base specification is the “true” scenario, the greatest performance loss (9%) comes from mistakenly applying the high initial infections policy.

We find that performance costs in percentage terms from applying the wrong policy from this set are typically modest (low single digits) albeit with notable exceptions. For example, when the “truth” is that we have a weak vaccine for ages 60+, several policies applied perform very poorly relative to the true optimal policy since they substitute vaccine away from younger essential workers to ages 75+. A few of the policies were generally less robust across various true models, specifically those for high initial infections, strong NPI, and weak vaccine 60+. The Base scenario policy performed reasonably well across true alternative models, with the largest loss arising (7%) when children are not less susceptible (even susceptibility).

Equivalent versions of Fig. 4B for minimizing deaths or infections are provided in SI Appendix C. When the focus is minimizing deaths, the pattern of performance between scenarios is very consistent with YLL in Fig. 4.

However, the scope for performance loss is larger overall–up from a maximum of 26% for YLL to 46% for deaths. When the focus is infections, the range of performance loss is much less intense at 7%. For infections, this relatively robust performance arises because optimal policies are much more similar across scenarios when minimizing infections (compared to the other objectives). Given greater scenario-driven heterogeneity in policies for minimizing YLL or deaths, there is greater opportunity for performance loss from specification error.

#### 2.1.2 A gradient over four key parameters

For further sensitivity analysis, as shown in Fig. 5, we assessed how optimal vaccine allocation policy changed along a gradient for four key model inputs: non-social distancing NPI effectiveness (e.g. mask wearing) which determines the initial reproductive number (when the vaccine first becomes available); initial infections; monthly rate of vaccine supply; and vaccine effectiveness.

**Figure 5:**
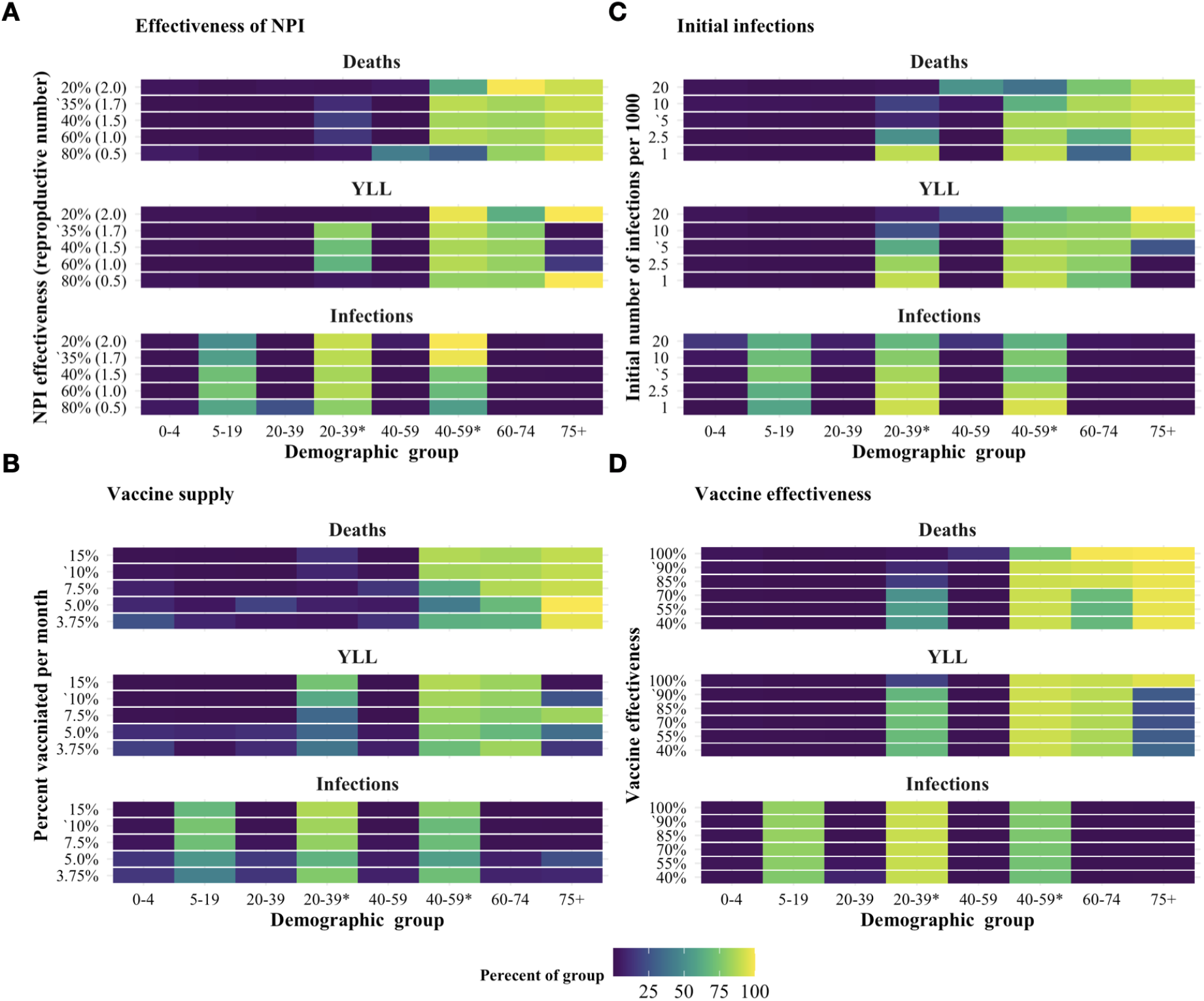
The total percent of each demographic group vaccinated after 3 months under the optimal dynamic policy. Each panel shows the effect of varying a key parameter relative to the Base model: (A) effectiveness of NPI, which determines the initial reproductive number (when the vaccine first becomes available); (B) monthly rate of vaccine supply; (C) initial infections; and (D) vaccine effectiveness. Base scenario parameter values are indicated with an apostrophe (‘).

Echoing sensitivity results reported above, variation in these parameters had little effect on the optimal policy for minimizing infections. But we found systematic differences in the policies for minimizing YLL and deaths. Essential workers, ages 60-74 and ages 75+ remained the highest priority groups across the full range of parameters tested but there was substitution between younger essential workers (20-39^*^) and the older age groups.

In most instances the percent of vaccines responded in relatively monotonic fashion as parameters varied. For example, consider the objective of minimizing deaths. As depicted in Fig. 5A:D, prioritization of essential workers fell and 60+ or 75+ increased as (1) initial infections grow; (2) vaccine supply decreases; or (3) vaccine effectiveness increases. In a few instances, the percent of vaccine allocated to a given group responded non-monotonically to variation in the parameter. For example, for effectiveness of NPI in Fig. 5A, the allocation skewed towards 75+ and away from essential workers when the parameter was very high and very low.

These results indicate that when focusing on deaths or YLL, if transmission cannot be reduced quickly by the vaccine—due to limited supply, high reproductive numbers or large initial number of infections—typically this initial supply is most efficiently used to directly protect individuals with the greatest risk of death if infected. This pattern differs for vaccine effectiveness: we find that as the effectiveness of the vaccine decreased, supply is substituted away from the older (higher risk) age groups to essential workers. This difference is consistent with the fact that as vaccines become less effective for a given individual, protecting vulnerable individuals is better achieved by reducing population-level transmission.

#### 2.1.3 Changes to model structures

As a final sensitivity analysis, we examined robustness of the results to three alternative model structures: (1) clustered essential workers, where essential workers only contact other essential workers in the workplace, (2) concentrated essential workers, where relative to the Baseline scenario, the portion of the working age population deemed “essential” is half (20%) and they have approximately double the contact rate; and (3) leaky vaccine, where rather than working perfectly for 90% of individuals, vaccinated individuals have reduced susceptibility to infection, infectiousness and risk of death if infected. A more detailed discussion of these models is included in SI Appendix D.

We found that the qualitative nature of the solutions remained constant across each of these alternative models, with some minor differences. Treating the essential worker group as a cluster increased the proportion of vaccine allocated to ages 60+ when deaths and years of life lost were considered. This shows that when essential worker contacts are clustered within-group, this reduces the indirect protection that vaccinating these individuals provided to others. Conversely, concentrating the essential worker group (to a more select group with higher contact rates) increased the fraction of these individuals vaccinated. This shows that select essential workers with especially high contact rates (e.g. medical professionals and essential retail workers) are particularly strong candidates for early vaccination.

## 3 Discussion

Key insights and results from our analysis are summarized in Box 1. Together these lessons show the strong implications of considering dynamic solutions, social distancing and essential workers (given their limitations in social distancing) for vaccine prioritization.

### Box 1.

**Key insights and results**

1. **Benefits:** Prioritization can reduce a particular undesirable outcomes (deaths, YLL, or infections), by 32-40% in the Base scenario (or 17-44% depending on the alternative scenario).
2. **Objectives:** Moving from minimizing infections to YLL to deaths, boosts each of the following— benefits from vaccination targeting, prioritization differences between scenarios, and (therefore) the sensitivity of optimal prioritization to scenario.
3. **Essential workers:** Essential workers are a high priory group across all three objectives (deaths, YLL and infections). Policies allowing for targeting based on essential worker status substantially outperformed those that consider age only.
4. **Dynamic prioritization:** Dynamic prioritization (1) is responsive to the initial and evolving disease status, and (2) generates substantial improvement in outcomes relative to a static prioritization. Diminishing marginal returns to additional vaccination within a group drives a shift to other groups before 100% vaccination of the first group is achieved.
5. **Trade offs:** There are substantial trade offs between the three objectives we considered. For example, policies that minimize deaths forgo most of the opportunity to reduce infections (though minimizing YLL involves much more modest trade offs). These trade offs are typically stronger when policies do not allow for targeting based on essential worker status.
6. **Widening prioritization:** As vaccination rates rise, precise prioritization becomes less critical and targeting widens to a larger set of groups.
7. **Sensitivity:** The high priority groups remain consistent across the range of parameters considered. However, minimizing deaths or YLL, the fraction of vaccine allocated to essential workers and ages 60+ depends on: the number of infections and reproductive number when the vaccine became available; the supply of vaccines; and vaccine effectiveness.

Our analysis of COVID-19 vaccine prioritization uniquely accounts for two critical needs: (1) dynamic prioritization given gradual roll out of vaccine during an active pandemic, and (2) attending to significant heterogeneities in work contacts among the adult population due to the ability of many to work from home. These two novel features significantly change optimal vaccine prioritization. Given gradual vaccine deployment, static policies are out-performed by dynamic polices, which narrowly target a small number of demographic groups and (after substantial coverage of them) switch to lower priority groups. Static policies identify a set of high priority groups but not how to order them when phased deployment is necessary. More significantly, targeting essential workers (or other adults with large number of work contacts) significantly reduces not just the adverse outcome of focus but also trade offs for remaining outcomes. For example, when minimizing deaths, allocation that differentiates essential workers substantial lessens the degree to which infections and YLL climb from the minimum achieved when each is optimized on its own.

Notable existing analysis of optimal COVID-19 vaccination targeting in preprint form includes Matrajt et al. (15), Bubar et al. (16) and Hogan et al. (17). Before comparing and contrasting results some key modeling differences should be noted. These preprints consider a wider range of vaccine availability than considered here. Our analysis uniquely incorporates non-pharmaceutical interventions (NPI), including social distancing and non-social distancing (e.g. mask wearing). Doing so allows us to account for differences between groups like essential workers constrained in distancing versus others who are much less so. All three preprints implement static optimization where vaccines are allocated and administered in a one-shot process. Our allocation is dynamic, responding to changing epidemiological conditions over a six-month period. Finally, all three model vaccines as “leaky”, i.e., reducing the probability that a susceptible individual will be infected (and the probability of severe disease (17)). Bubar et al. also considers an “all-or-nothing” vaccine that is 100% effective for a fraction of the population. In our Base model the vaccine is “all-or-nothing”, though we also consider a leaky vaccine, as discussed at the end of the Results.

Matrajt et al. (15) found that optimal strategies to minimize deaths and years of life lost will either exclusively target groups with high infection fatality rates maximizing the direct benefit of vaccines, or will target groups with high rates of infection maximizing the indirect benefits of the vaccine. In contrast, our results indicate that optimal policies initially target groups with high risk of infection and then switch to targeting groups with high infection fatality. This difference most likely follows from our dynamic versus static allocation. The switching behavior we identify is consistent with past work on pandemic influenza vaccine prioritization, which suggests that early in an outbreak when the infection rate is growing targeting spread (maximizing indirect benefits) is more efficient, but later when the infection rate is leveling off or declining, maximizing direct protection is most efficient (20).

Bubar et al. found that prioritizing adults older than 60 years of age is a robust strategy for minimizing deaths. In contrast we find that working-age adults are a key priority group, particularly older essential workers. These differences may either arise from our allowance for social distancing and/or dynamic allocation. Our accounting for social distancing on COVID-19 transmission increases the modeled benefits of targeting essential workers, who are less able to substantially reduce their social contacts than individuals over 60. Furthermore, as discussed above, the ability of dynamic policies to switch over time allows the allocation schemes we discuss to capture the benefits of using the initial vaccine supply to slow transmission without sacrificing direct protection of more vulnerable individuals later on.

Although this model provides useful insight for the policy-making process, a number of caveats are in order. In reality the risk of infection varies continuously across individuals, even between different “essential” occupations. While our model is unique in capturing differences between essential and non-essential workers, the representation of these differences is simplified by averaging the total number of contacts over a group with high work contacts (essential workers) and a group with lower rates of work contacts. This allows us to demonstrate the importance of this heterogeneity in the adult population relative to the standard age-only models, indicating that policy makers should strongly consider occupation-differentiated vaccine allocation strategies.

While we explored a large set of alternative scenarios, further extensions remain for future work. For example, if certain groups (e.g., children or seniors) experience significant vaccination side effects, prioritization might shift away from these groups (22). From a logistical perspective, vaccination will occur through various points of contact with the community (pharmacies, clinics, schools, etc.). Constraints imposed by the distribution network used will affect the relative costs of reaching various subgroups.

From a behavioral perspective, vaccine hesitancy may influence the ability to achieve vaccination priorities, especially as coverage of the population increases. In general, we find that it is not necessary or even ideal to vaccinate all of the susceptible individuals in a demographic group, at least given the level of 60% of the population vaccinated considered here. Thus, at least initially, some level of vaccine hesitancy may have limited material impact. However, hesitancy may play a more significant role in the longer run, especially if hesitancy rates are large and herd immunity proves difficult to achieve (e.g. if vaccine effectiveness is low, and/or NPI relaxation is aggressive). Vaccine hesitancy that is concentrated in a particular community or demographic group could also change the optimal prioritization strategy. Similarly, adjustments would be needed if groups differ in the duration of vaccine effectiveness or diligence in obtaining a second dose of the vaccine where (and when) necessary.

For simplicity we limited policy objectives to a set of concise metrics of health outcomes (minimizing expected cases, YLL, or deaths). However, other health-related metrics such as protecting the most vulnerable and social values such as returning to school, work and social life are important to consider. Our analysis reveals that optimal strategies for minimizing deaths and YLL are broadly aligned with the goal of protecting the most vulnerable. These solutions target essential workers who are the least able to participate in NPI such as social distancing and thus are the most at risk of infection, and individuals over the age of 60 yrs. who have the highest risk of death if infected by the disease. Other social values such as returning to school will most likely change the allocation schemes to offset the risk created by relaxing social distancing. For example, if allowing children to return to school was a high priority, then allocation strategies might be tilted towards targeting school-age children and teachers. A detailed analysis of optimal vaccine allocation given the relaxation of social distancing to achieve particular social objectives is a key direction for future research.

## 4 Methods

### 4.1 Model

To investigate the impact of vaccination strategies on the COVID-19 pandemic in the U.S., we employed a structured compartmental transmission model similar to (23). We incorporated the demographic structure of the population by tracking six age groups in the set *J* = {0-4, 5-19, 20-39, 40-59, 60-74, 75+}. We then extend this set to differentiate essential workers by splitting the two prime working age groups into two groups—non-essential workers (20-39, 40-59) and essential workers (20-39*, 40-59*)—yielding four groups of prime working age individuals and a total of eight demographic groups in *J* = {0-4, 5-19, 20-39, 20-39^*^, 40-59, 40-59^*^, 60-74, 75+}. For each demographic group we tracked 9 epidemiological states: susceptible (*S*), protected by a vaccine (*P*), vaccinated but unprotected (*F*), exposed (*E*), pre-symptomatic (*I*_*pre*_), symptomatic (*I*_*sym*_), asymptomatic (*I*_*asym*_), recovered (*R*) and deceased (*D*). In Fig. 1 we display the compartmental diagram and directions of transitions between epidemiological states.

We modeled the COVID-19 transmission dynamics using a system of coupled ordinary differential equations for each demographic group, indexed by *i* and *j*. The transmission rate was given by the product of the transmission probability (*q*), the age-specific susceptibility (*s*_*i*_), strength of non-pharmaceutical interventions (*θ*), the relative infectiousness of each symptom type (*τ*_*m*_)—where *m* ∈ *M ≡ {asym, pre, sym}*—and the rate of contact (*r*_*m,i,j*_) between infected individuals with symptom type *m* from group *j* and susceptible individuals from group *i*. The exogenously given population vaccination rate at time *t* is given by *v*(*t*), where units of time are days.^4^ In our Base model we assume that for each individual the vaccine either works or it does not (though we also consider vaccines that are partially effective for all vaccinated in our sensitivity analysis). Individuals in group *i* are vaccinated at a rate of *µ*_*i*_*v*(*t*) and a fraction of the those (∈_*i*_) are protected while a fraction remain susceptible and move to the failed vaccination category (*F*).^5^ Once infected, individuals move from exposed to pre-symptomatic at rate 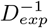. Pre-symptomatic individuals become symptomatic or asymptomatic at rates *σ*_*asym*_*/D*_*pre*_ and (1 − *σ*_*asym*_)*/D*_*pre*_ respectively. Asymptomatic individuals recover at an uniform rate 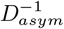 and symptomatic individuals either recover or die at a rate of (1 − *δ*_*a*_)*/D*_*sym*_ or *δ*_*a*_*/D*_*sym*_, respectively, where *δ*_*a*_ is the age-specific infection fatality rate. These assumptions yield the system of differential equations described in SI Appendix A.1, with parameter values given in SI Appendices A.2 and A.3.

### 4.2 Contact rates

Contact rates indicating the level of direct interaction of individuals within and between groups drive the transmission dynamics in the model. We built the contact matrices used in this model from the contact matrices estimated for the U.S. in (13). These estimates are given for age groups with five year age increments from 0 to 80 yrs. These estimates were aggregated to provide estimates for the six-level age structure used in our model. We also extended these data to estimate the contact rates of essential workers. A detailed derivation of these contact rates can be found in SI Appendix A.6. In short, we assumed that essential workers have on average the same pattern of contacts as an average worker in the population in the absence of social distancing. We then scaled the contact rates for essential and non-essential workers to represent the effects of social distancing and calculated the resulting mixing patterns assuming homogeneity between these groups.

We constructed contact matrices for each of four locations, *x* ∈ *{home, work, school, other}*, following (13). The total contact rate for an asymptomatic individual before the onset of the pandemic is given by the sum of these location specific matrices. However, it is clear that populations are exhibiting social distancing in response to the pandemic (24). We further expect symptomatic individuals to change their behavior in response to the illness. We account for these behavioral changes as described next.

### 4.3 Social distancing

Expression of symptoms and social distancing policies are likely to change individuals’ behaviors over time. To model these changes we scaled the contribution of each contact matrix for location *x*:

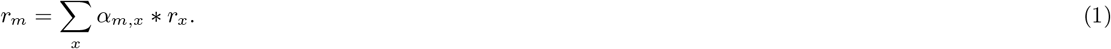

The weights *α*_*m,x*_ depend on disease and symptom status (*m*) and location (*x*) as specified in Table 2. We scaled social contacts for symptomatic individuals following changes in behavior observed among symptomatic individuals during the 2009 A/H1N1 pandemic (25). For those without symptoms (susceptible and asymptomatic) the weights were specified to match reduced levels of social contacts as the product of social distancing policies. Home contact rates were held constant, and non-household contact rates were roughly based on survey data from (14). However, levels of social distancing have varied significantly over time and between locations. To account for this variability we tested a range of alternative levels in addition to the Base model. The results for these alternative parameter values are discussed in appendix D.2. Also, notably we do not consider the seasonality of contact rates for children in the scenarios where schools are modeled as closed. This would likely have limited impact on the optimal solutions, but when this is not the case we may over or underestimate the importance of school contacts depending on the time of year when vaccines are distributed.

**Table 2:** Weights on contact rates for a given disease and symptom type (*m*) and location/activity (*x*) under social distancing. When essential and non-essential-worker weights are both needed the former is marked with a star.

| Disease and symptom type | Contact rate weights, $\alpha_{m,x}$ | | | |
| --- | --- | --- | --- | --- |
|  | *home* | *work* | *school* | *other* |
| symptomatic | 1.0 | 0.036 | 0.036 | 0.075 |
| susceptible or asymptomatic | 1.0 | 0.4*, 0.1 | 0.3 | 0.4 |

The proportion of essential workers in the population was set to be consistent with estimates of the portion of jobs that can be done from home (26) and estimates from the U.S. Cyber-security and Infrastructure Security Agency, which indicate that 70% of the workforce is involved in these essential activities (e.g. healthcare, telecommunications, information technology systems, defense, food and agriculture, transportation and logistics, energy, water, public works and public safety) (27). However, essential workers are not a cleanly defined group of individuals and there is significant heterogeneity in the level of contact rates within this group. As a robustness check on this Base scenario approach, we also tested a model with a smaller number of essential workers with higher contact rates. Results from this model are discussed in SI Appendix D.

### 4.4 Transmission rate and vaccine effectiveness

The model was calibrated to match the predicted *R*_0_ for COVID-19 in the U.S. (see SI Appendix Table 3) by solving for probability of transmission *q*, assuming a naive (pre-pandemic) population. Details of this procedure are provided in SI Appendix A.5.

In our Base model we considered vaccine effectiveness of 90%. This level is at the low end of the range of estimates reported (90-95%) for reduction in symptomatic infections in the fall of 2020 from phase three clinical trials (28). We selected the low end since real world performance is typically somewhat lower than clinical trial effectiveness, e.g. due to imperfect implementation of dual-dose timetables and or cold storage requirements. We also assume this effectiveness is the same across age groups since initial evidence does not show significant differences between subgroups (29). As an alternative, lower-bound scenario we considered vaccine effectiveness of 50% since this is the minimum expectation of the U.S. FDA for approval (30). Finally, we considered a case where the vaccine is less effective for ages 60+. The phase three trials do not fully resolve the effectiveness of the vaccines by age, leading to uncertainty. This scenario represents a worst case scenario where the vaccine is much less efficacious for the most sensitive groups.

### 4.5 Initial conditions

Because the expected epidemiological conditions *{I*_*pre*_(0), *I*_*asym*_(0), *I*_*sym*_(0), *S*(0)} by the time the initial vaccine doses are ready for deployment are uncertain, we consider a range of possible values from 1 case per thousand to 20 cases per thousand. These cases were apportioned between demographic groups to reflect the attack rates of COVID-19 for each group under the given social distancing policy. Alternative levels considered for initial conditions are described in SI Appendix A.4 and appear in Results (see Table 1, Fig. 4 and Fig. 5C).

### 4.6 Vaccine prioritization optimization

The planner’s decision problem is to allocate the daily supply of vaccine (*v*(*t*)) across the demographic groups according to a given objective. We assume that this group allocation vector, *µ*, can be chosen on a monthly basis at the beginning of each of the first six decision periods. We also assume that only susceptible individuals are vaccinated. We numerically solved for vaccine allocation strategies that minimize the total burden associated with three different health metrics: deaths, years of life lost (YLL) or symptomatic infections:

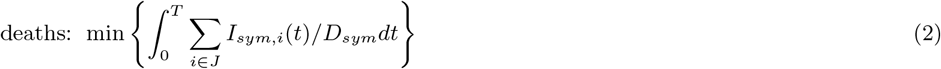

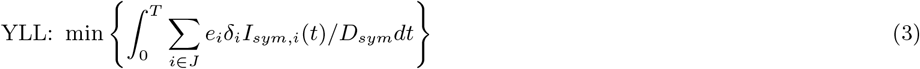

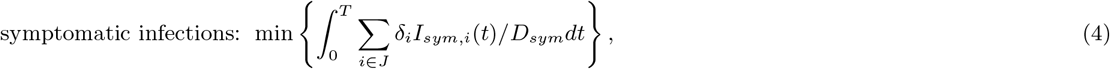

where *e*_*i*_ is the years remaining of life expectancy for group *i* and with a six month time horizon (*T* = 180 days). Preventing deaths and years of life lost are “consensus value(s) across expert reports” (4, p. 2052) while some argue that “protecting public health during the COVID-19 pandemic requires…minimizing COVID-19 infection” (5, p. 10).

We solved for the optimal allocation of available vaccines across demographic groups for each month over six months. We identified the optimal solution using a two-step algorithm. In the first step we used a genetic algorithm similar to (31) to identify an approximate solution. This approach uses random sampling of the potential solution space to broadly explore in order to avoid narrowing to a local and not global minimum. In the second step we used simulated annealing to identify the solution with precision. At a given optimal solution, it may or may not be the case that the outcome of interest (e.g. minimizing deaths) is sensitive to small changes in the allocation decision. Thus, around the optimal allocation we also identified nearby allocations that produce outcomes that are less desirable but still within 0.5% of the optimized outcome. A detailed description of the algorithm is given in SI Appendix A.7. All code for the optimization was written in the Julia programming language (32).

To assess the benefits of (1) using a dynamic allocation policy and (2) differentiating by essential worker status in addition to age, we constructed two constrained policies: a static policy and an age-only policy. The static policy was found by allowing the proportion of vaccine allocated to each age group to be chosen once when the vaccine first becomes available and then applied constantly over time.^6^ The age-only policy simply involves constraining allocation choices age groups (not differentiated by essential worker status)—vaccines allocated to working age groups accrue to essential workers simply in proportion to their relative share of these groups.

### Data access

All data used for informing the numerical analysis are freely available at the source noted for each measure. The data and code used to initialize and run the models will be made available on Github upon publication, https://github.com/XXX.

## Data Availability

All data used in the article are publicly available from the relevant citation included in the text. Code for the analysis may be obtained from the authors upon request.

## Acknowledgements

This material is based upon work supported by the National Science Foundation (NSF) Graduate Research Fellowship Program under Grant No. 1650042. Any opinions, findings, and conclusions or recommendations expressed in this material are those of the authors and do not necessarily reflect the views of the NSF. MS is supported by an Emergency COVID-19 Seed Award from the California Breast Cancer Research Program of the University of California, Grant No. R00RG2419. GC is partially supported by NSF grant No.’s 2026797, 2034003, and NIH R01 GM 130900. For helpful comments, we thank participants in the MIDAS network COVID-19 modeling seminar and the World Health Organization Strategic Advisory Group of Experts on Immunization Working Group on COVID-19 Vaccines (Impact Modelling subgroup).

## Appendix

### A Model specification, parameterization and optimization

#### A.1 Model dynamic equations

The dynamic equations specifying transitions between the disease states are as follows:

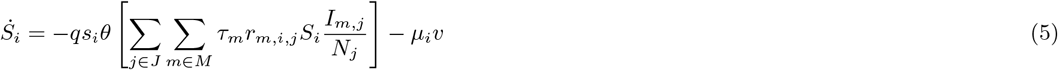

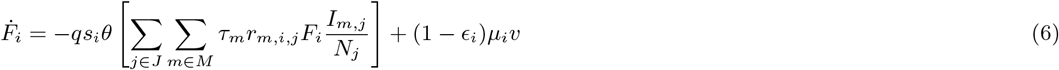

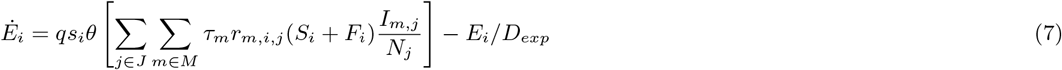

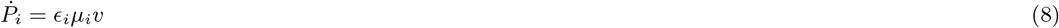

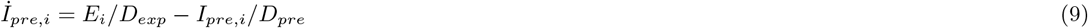

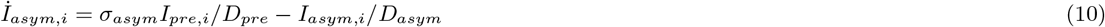

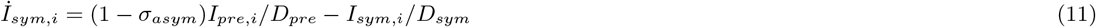

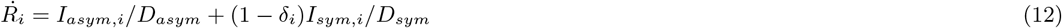

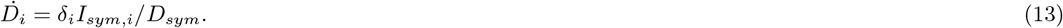

To reduce clutter we have suppressed the time index *t* on each of the state variables, the vaccine allocation vector *µ*_*i*_, and the vaccination rate *v*.

#### A.2 Model parameters

#### A.3 Parameters for alternative scenarios

#### A.4 Initial conditions

The number of susceptible, infected and recovered individuals is likely to vary by region and will depend on the time when the vaccine becomes available. Because of the likely variation in an uncertainty about this parameter we test a range of values from 1 symptomatic case per 1000 when the vaccine become available to 20 cases per 1000. The infections are assumed to be distributed between groups in accordance with the stable distribution of cases when the epidemic is growing exponentially. The portion of each group infected at time *t* = 0 in the Base parameter set is given below.

##### Calibration

The relationship between the basic reproduction number, *R*_0_, and parameters governing transmission and epidemiological characteristics is given by the so-called next-generation matrix:

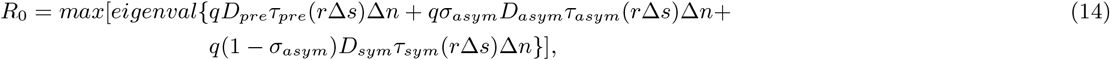

where the maximum eigenvalue operator wraps several terms including *r*, the social contact matrix, *s*, the age-specific susceptibility rate, *n*, a vector of the proportions of the population in each demographic group and Δ, an operator that signifies multiplying each row of a matrix by the corresponding entry in the vector. For symptom type *m* ∈ *{asym, pre, sym}*, the constants *D*_*m*_, *τ*_*m*_ and *σ*_*m*_ represent the duration, relative infectiousness of an individual and the probability of type *m*, respectively.

We first set a baseline *R*_0_ = 2.5 as estimated by (37). We then solve for the transmission probability parameter, *q*, using Equation 14, assuming a naive (pre-pandemic) population. We then scaled *q* by a fixed factor *θ* ∈ [0, 1] to reflect the impact of non-pharmaceutical interventions (NPI) like masks, hand washing and maintaining distance when contacts are made.

#### A.6 Contact matrices distinguishing essential workers

Estimated contact rates for the U.S. were obtained from (13) who used population-based contact diaries from the European POLYMOD survey to project to other countries, including the U.S. These included contact rates for 16 age classes in five year increments from ages 0 to 80. We collapsed these to five age groups (0-4, 5-19, 20-39, 40-59, 60-80) using population-weighted sums:

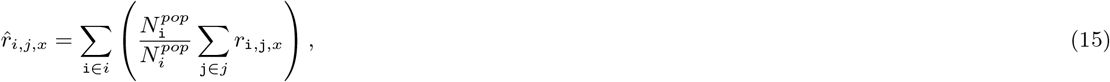

where `{i, j}` are the subscripts for the five year age bins, *{i, j}* are the subscripts for the larger age bins, *r*_i,j,*x*_ is the average number of daily contacts a person in group i makes with a person in group j for activity/location *x*, and 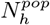 is the population size for age group *h*.

The total number of *i*-to-*j* contacts must equal the total number of *j*-to-*i* contacts: 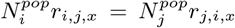. Because numerical issues—estimation in (13), bin discretization and rounding—can lead to small differences, we ensure this condition holds by imposing,

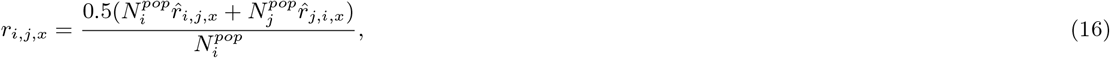

where the numerator is the mean of the two measures of total contacts between groups *i* and *j* and the denominator transforms the result to per-capita in *i*.

Setting essential worker contact rates requires additional assumptions and attention to the activity/location. We define the essential worker indicators *e* ∈ *{n, y}* for “no” and “yes”. Our grouping is such that all essential workers (*e* = *y*) are employed but non-essential-workers (*e* = *n*) are a mix of employed and not employed. Let *e′* represent the indicator for a second group which can be equal or not equal to the value for *e*.

In the case of all activities/locations *x* that are not *work*, contact rates are given by

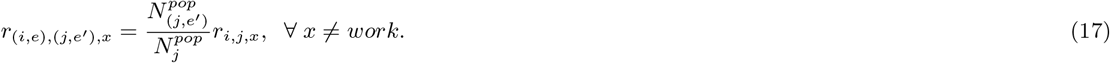

This follows from the assumption that contacts made by any group (*i, e*) with any other group (*j, e*′) are independent of *i*’s essential worker status. Thus, we only need to split contacts *r*_*i,j,x*_ into those made with essential worker type *e*′ = *y* versus the remainder with type *e*′ = *n*, i.e. given the share 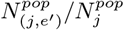.

Estimating contacts when *x* = *work* involves a larger number of steps. We first address contacts made by essential workers (*e* = *y*) before turning to non-essential workers (*e* = *n*). For *e* = *y*, let the share of the working age population (20 − 59) in group *i* that is employed be given by *p*_*i*_.

We assume that all of the work contacts are attributable to employed adults resulting in an employed adult contact rate of *r*_*i,j,work*_*/p*_*i*_. Then the contact rate of essential workers (*e* = *y*) in group *i* with age group *j* is

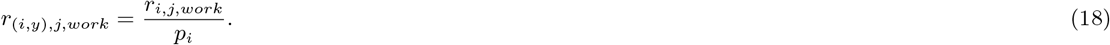

Let the fraction of working age group *i* that is employed in an essential worker role be given by *p*_*i,y*_. The average workplace contact rate for non-essential-workers in group *i* with group *j* is given by

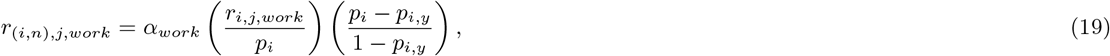

where *α*_*work*_ *<* 1 scales for social distancing and the final term in brackets scales for the share of non-essential workers that are employed and thus have contacts at *work*.

Finally, we assume that the average *work*place contact rate for an individual of type (*i, e*) with individuals of type (*j, e*′) is given by the partial contact rate *r*_(*i,e*),*j,work*_ times the proportion of total work contacts of individuals in group *j* that are made by individuals in sub group *e*′:

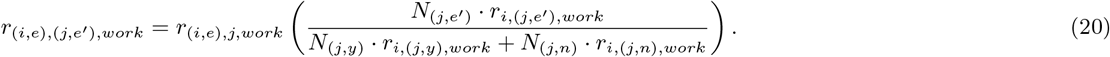

In addition to The formulation described above we also considered a scenario where essential workers contacts were clustered (i.e. individuals only contact others of the same essential worker status at work). The work contact rates for each group are calculated in the same manner as described above, but we assume that the c work contacts between essential and non-essential workers are zero.

These two model formulation represent two extremes. Work contacts are likely to be concentrated among others of the same essential status (as opposed to formulation one) but essential workers are likely to have some contacts non-essential workers in the work place.

Finally we scale the work contacts for age groups that are not separated into essential and non-essential workers (5-19, 60-80) to match the scaling for prime working age classes.

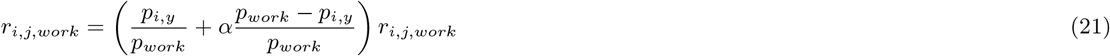

#### A.7 Optimization algorithm

The optimization algorithm used in our analysis is split into two parts. First a genetic algorithm is run to identify an effective strategy near a global optimum. This solution is then refined via simulated annealing.

Genetic algorithms take inspiration from the natural process of evolution, and work by randomly sampling a populating of candidate solutions, selecting a set of survivors based on the candidates performance against the objective function, information from these survivors is then used to generate a new generation of candidates solutions, and so forth (31). The genetic algorithm executes the following steps:

1. Sample *N*_*t*=0_ candidate solutions *{x*_*n,t*=0_} from a Dirichlet distribution with parameter *α*_0_.
2. Each candidate solution is evaluated with the objective function.
3. The bests *K*_*t*=0_ candidates 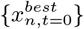 are solved and the distributions parameter *α*_0_ is updated to *α*_1_ which is the mean of 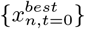 times the entropy parameter for that time step *η*_*t*_. The entropy parameter determines how concentrated new samples will be around the mean of the selected samples in the prior step.
4. Steps 1 to 3 are repeated for a fixed number of iterations *T* and the best candidate solution sampled at any iteration is returned. The values *N*_*t*_, *K*_*t*_ and *η*_*t*_ are tuned for each step to maximize performance.

Simulated annealing is based on thermodynamic models of cooling metals. Briefly, the algorithm is initialized by sampling a candidate solution *x*_0_, this candidate solution is updated by sampling a new candidate solution *x*_*t*_ from a proposal distribution centered around *x*_0_. This solution is either accepted and replaces the current *x*_0_ or it is rejected and a new candidate solution is drawn using the existing value of *x*_0_. The proposed solutions *x*_*t*_ are accepted if they perform better against the objective than the incumbent *x*_0_, if *x*_*t*_ > *x*_0_ it is selected with probability *p* = *exp* [−(*x*_*t*_ − *x*_0_)*/T*_*t*_]. large values of *T*_*t*_ increase the probability that a new candidate solution will be accepted allowing the algorithm to explore the solution space and move away from local minima. *T*_*t*_ is reduced over time to allow the algorithm to start exploring the solution space and then eventually stabilize on a global minimum. The simulated annealing executes the following steps:

1. Initialize a chain with value *x*_0_. Generate a new sample from the proposal distribution *x*_*t*_ ∼ *tr*(*N* (*tr*^−1^(*x*_0_), *σI*)) where the transform *tr* from **R**^*n*^ to the solution space. Initialize a counter *i* that tracks the number of iterations.
2. If *x*_*t*_ *< x*_0_ replace *x*_0_ with *x*_*t*_, update *i* = *i* + 1 and repeat from step 1.
3. If *x*_*t*_ > *x*_0_ sample *µ* ∼ *unif* (0, 1). If *µ* > *exp* (−(*x*_*t*_ − *x*_0_)*/T* (*i*)) then replace *x*_0_ with *x*_*t*_ update i and repeat from step 1. Otherwise save *x*_0_ and repeat from step 1. We used *T* (*i*) = *T*_0_*/i* as the temperature function.
4. Stop when *i* > *max iter*

These algorithms were tuned experimentally to consistently converge on a minimum solution on a test case. We used the minimum years of life lost under the Base parameter set as our test case. We found that numerical errors (defined as difference between the test runs) increased over the decision periods. This is caused by the face that the final decision periods are a region of very flat payoff, leading to a large number of solutions that perform very similarly but differ in this region

To quantify the sensitivity of the solutions to deviations in the outcome of interest, we sampled solutions near the optimal solution using a Markov chain. This algorithm is initialized at the optimal solution identified by simulated annealing and the genetic algorithm and samples are drawn from a proposal distribution and accepted if they perform within the desired tolerance (0.25%) of the optimal solution.

### B Static policies

We found that the patterns observed for our dynamic solutions were similar for the static policies. The static policies targeted the same high priority groups: ages 5-19 and essential workers when minimizing infections, and ages 60+ along with essential workers when minimizing deaths and YLL. Again consistent with the dynamic policies we found that optimal vaccine allocations were substituted towards essential workers when the effectiveness of the vaccine was low and when the reproductive number when the vaccine first became available was small. These patterns are shown in Fig. 7A. ^7^

**Figure 6:**
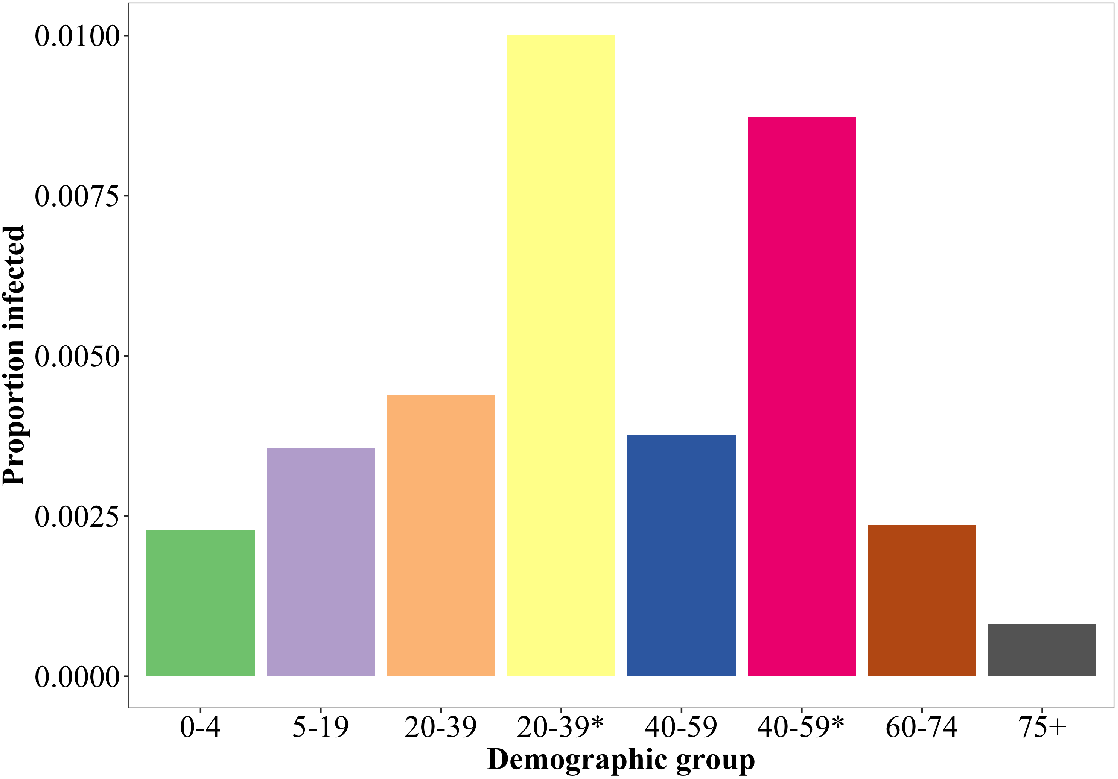
The proportion of each demographic group infected when vaccine distribution begins in the Base scenario.

**Figure 7:**
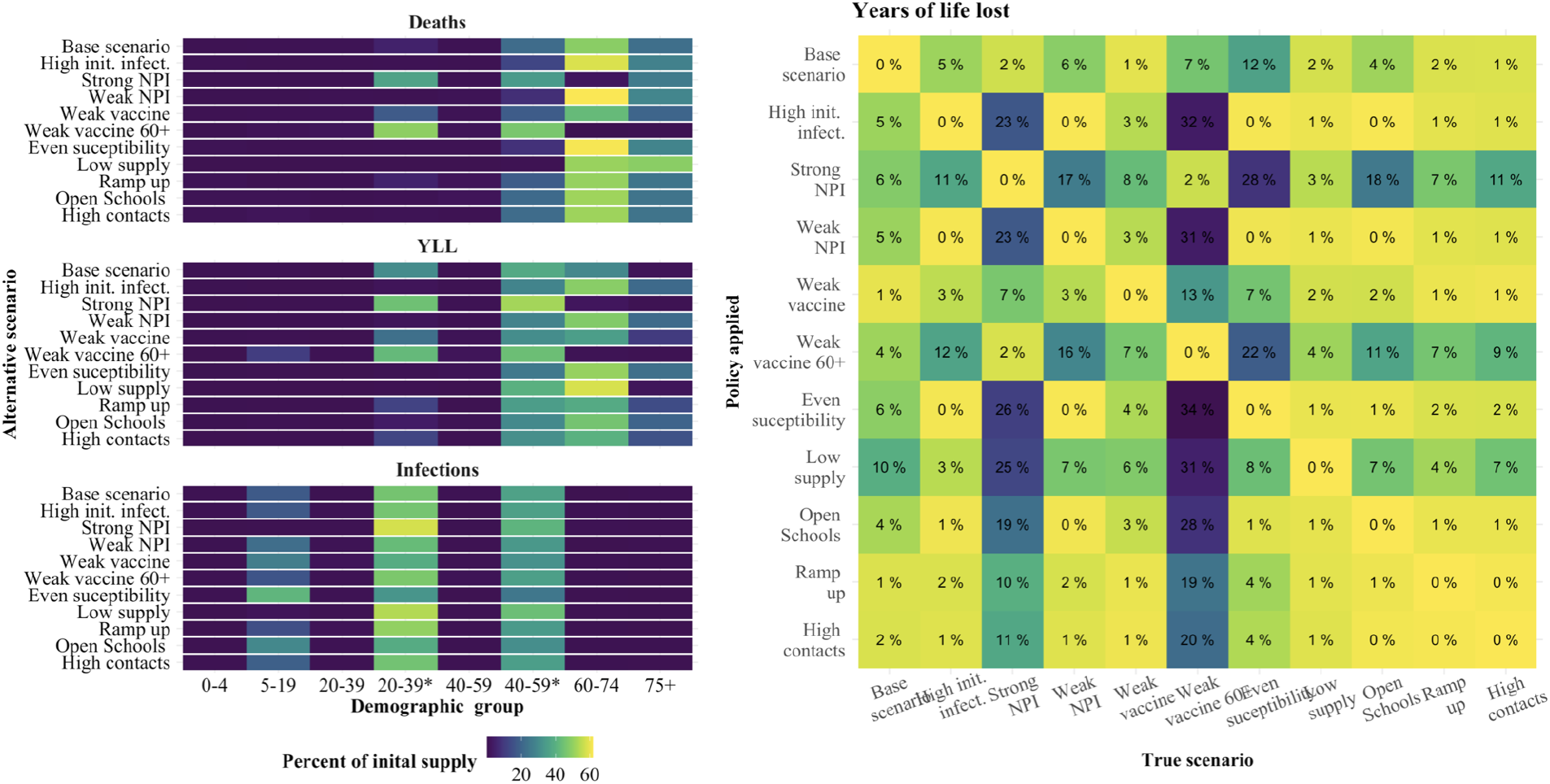
The sensitivity of static policies to the alternative scenarios as given by the percent of the initial supply allocated to each demographic group (A), the performance relative to the optimum allocation of each policy when applied to each of the alternative scenarios when the objective is YLL (B).

In Fig. 7B we show how robust the static policies are when applied to the “wrong” scenario. As with the dynamic policies we found that most polices performed very poorly when applied to the weak vaccine ages 60+ and the strong NPI scenarios, relative to the optimum. But, in general, deterioration in performance (due to a mismatch between the true scenario and the one driving the policy applied) was much worse for static than dynamic polices. One driver of this effect is that static polices identify one set of high priority groups and do not switch. Dynamic polices also identify one set of high priority groups but then also switch. Thus dynamic policies differ by when a group is prioritized, as opposed to the static polices which differ by which groups are a high (unchanging) priority.

### C Additional model robustness results

Here we present performance loss due to a mismatch between the true scenario and the one used to establish the allocation policy. Results are presented for either minimizing deaths (Fig. 8) or infections (Fig. 9).

**Figure 8:**
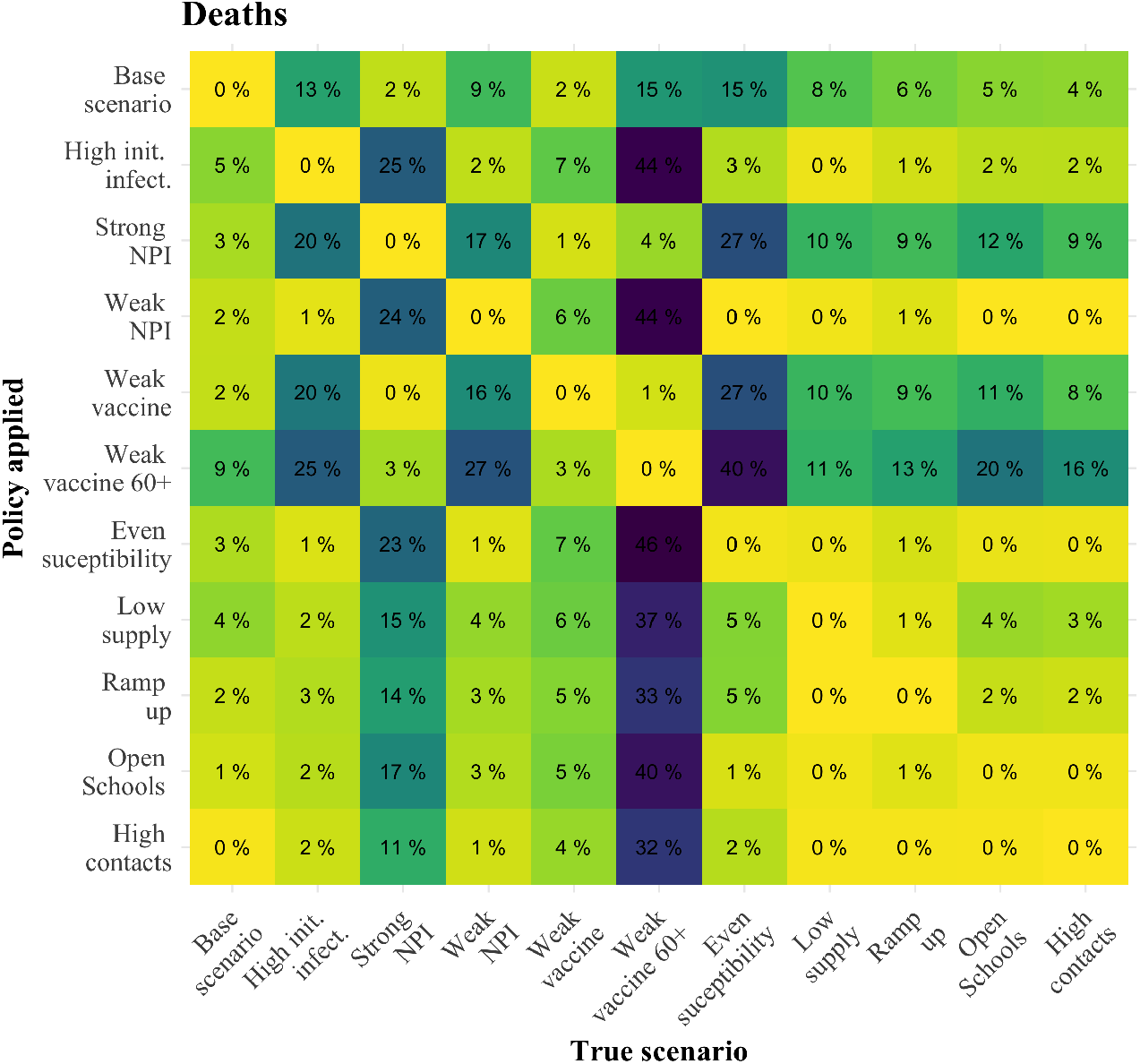
The percentage of additional deaths in excess of the optimum when applying a policy for an alternative scenario (row) to an alternative “true” scenario (column).

**Figure 9:**
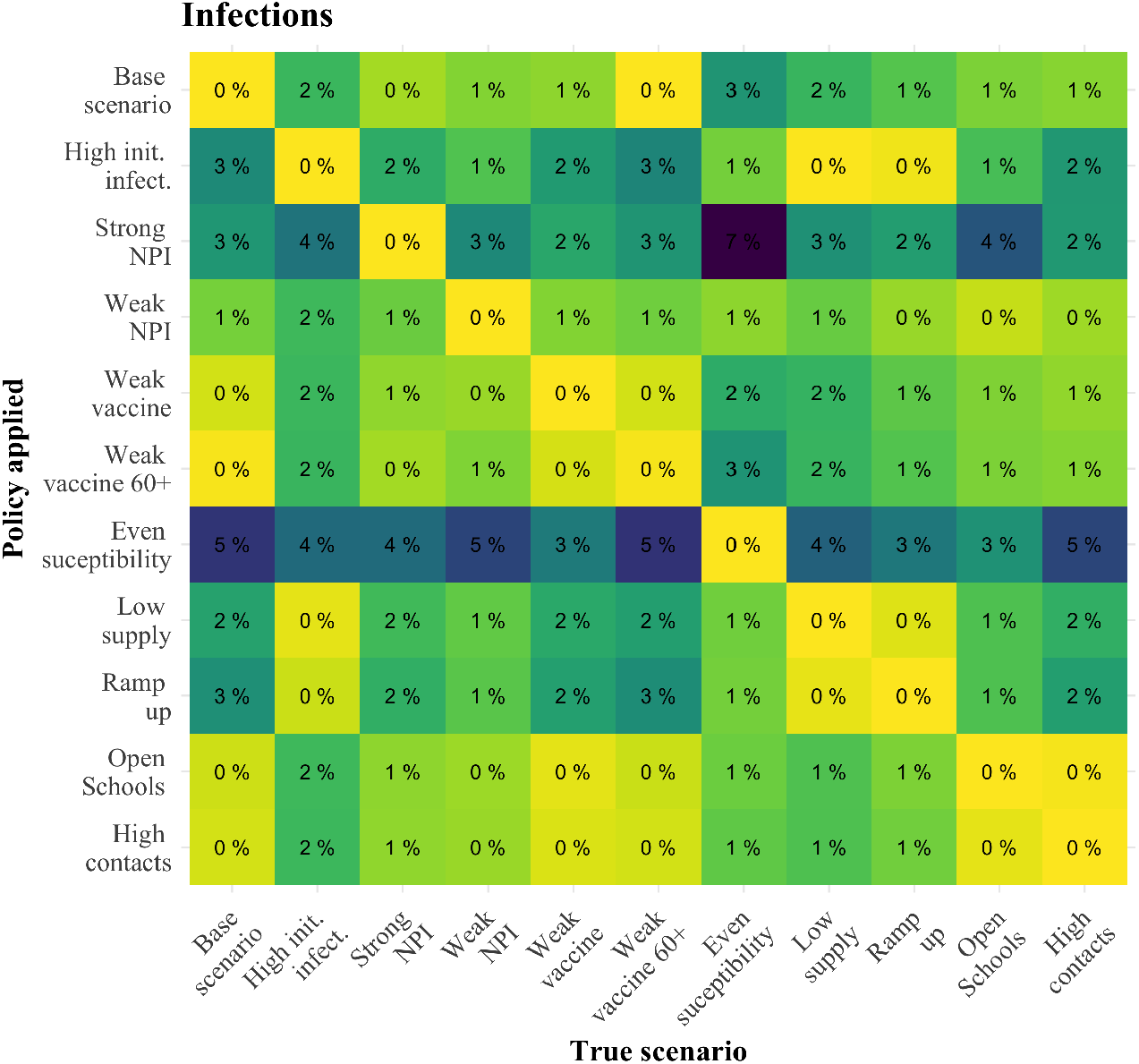
The percentage of additional infections in excess of the optimum when applying a policy for an alternative scenario (row) to an alternative “true” scenario (column).

### D Alternative model structures

In addition to considering a range of alternative parameter sets, we tested our results against three alternative model structures: (1) clustered essential workers: essential workers only contact other essential workers in the work place; (2) concentrated essential workers: 20% of the working age population are “essential” and have significantly higher (slightly above doubled) contact rates than essential workers in the Base model; and (3) leaky vaccine: the vaccine reduces the susceptibility of all vaccinated individuals to infection, and reduces their infectiousness and risk of death if infected. The corresponding parameter values for each alternative model are summarized below in Table 5 and the model structure for the leaky vaccine model is given in SI Appendix D.1. Results presented in Fig. 10 are discussed in the main text.

**Table 3:** Base model parameter values and sources.

| Parameter | Description | Base Value(s) | Source |
| --- | --- | --- | --- |
| $J$ | demographic groups: (1) age-only,<br>(2) age and essential workers | {0-4, 5-19, 20-39, 40-59, 60-74, 75+},<br>{0-4, 5-19, 20-39, 20-39*, 40-59, 40-59*, 60-74, 75+} | Assumed |
| $\sigma_{asym}$ | infection asymptomatic rate | 0.16 | (33) |
| $\delta$ | infection fatality rate (age-specific) | $\{6.7 \times 10^{-6}, 2.5 \times 10^{-5}, 0.0002, 0.002, 0.018, 0.12\}$ | (34) |
| $s$ | susceptibility (age-specific) | {0.5, 0.5, 1.0, 1.0, 1.0, 1.0} | (9) |
| $\tau_{pre}$<br>$\tau_{asym}$<br>$\tau_{sym}$ | relative infectiousness by symptom type | 0.51<br>0.51<br>1.0 | (23) |
| $D_{exp}$<br>$D_{pre}$<br>$D_{asym}$<br>$D_{sym}$ | symptom duration (days) | 3.0<br>3.2<br>3.5<br>7.0 | (23) |
| $\epsilon$ | vaccine effectiveness | 0.9 | Informed by initial COVID-19 vaccine effectiveness estimates (28) |
| $p$ | proportion of essential workers | 0.40 | Calculated with labor data (26, 35); alternative: (36) |
| $R_0$ | secondary infections in a naive population | 2.5 | (37), (38) |
| $q$ | transmission probability in a naive population | 0.053 | Calculated given $R_0$, $s$ and other parameters |
| $\theta$ | scaling factor for transmission probability due to NPI other than social distancing | 0.65 | Assumed (consistent with estimated COVID-19 $R_0$ under NPIs (38)) |
| $n$ | population shares: (1) age-only,<br>(2) age and essential workers | {0.06, 0.19, 0.27, 0.26, 0.19, 0.04},<br>{0.06, 0.19, 0.19, 0.08, 0.18, 0.8, 0.19, 0.04} | (39) |
| $e$ | remaining years of life expectancy (age-specific) | {76, 66, 50, 31, 17, 6} | (40) |
| $I_{pre}(0)$<br>$S(0)$<br>$R(0)$ | initial pre-sympt.<br>initial Susceptible<br>initial recovered | $0.005n$<br>$0.9n$<br>$0.08n$ | |
| $v$ | fraction of population vaccinated daily | 0.1/30 | Informed by comments from CDC Director to U.S. Senate Panel (41) |

**Table 4:** Parameter values that differ from the Base case for alternative scenarios.

| Scenario | Change from Base scenario parameters | Source |
| --- | --- | --- |
| High initial infections | 15 symptomatic infections per 1000 | Assumed |
| Strong NPI | $\theta = 0.5$ | Assumed |
| Weak NPI | $\theta = 0.75$ | consistent with 30-70% of U.S. population always wearing a mask (42) with 33-58% effectiveness (43) |
| Weak vaccine | $\epsilon_i \in \{0.5, 0.5, 0.5, 0.5, 0.5\}$ | Minimum value from FDA approval |
| Weak vaccine seniors | $\epsilon_i \in \{0.9, 0.9, 0.9, 0.9, 0.5, 0.5\}$ | Informed by influenza vaccine effectiveness |
| High susceptibility ages < 20 | $s_i \in \{1.0, 1.0, 1.0, 1.0, 1.0\}$ | Assumed |
| Low supply | $v(t) = 0.05/30$<br>allocation policies switched every 60 days | Assumed |
| Ramp up | $v(t) = \begin{cases} 0.05/30, & t \leq 60 \\ 0.10/30, & t > 60 \end{cases}$<br>Allocation polices switch for every 10% of population vaccinated | Informed by comments from the scientific head of the U.S. vaccine development program (21) |
| Open schools | $\alpha_{school} = 0.7$ | Assumed |
| High contacts | $\alpha_{social} = 0.5$ | Assumed |

**Table 5:**
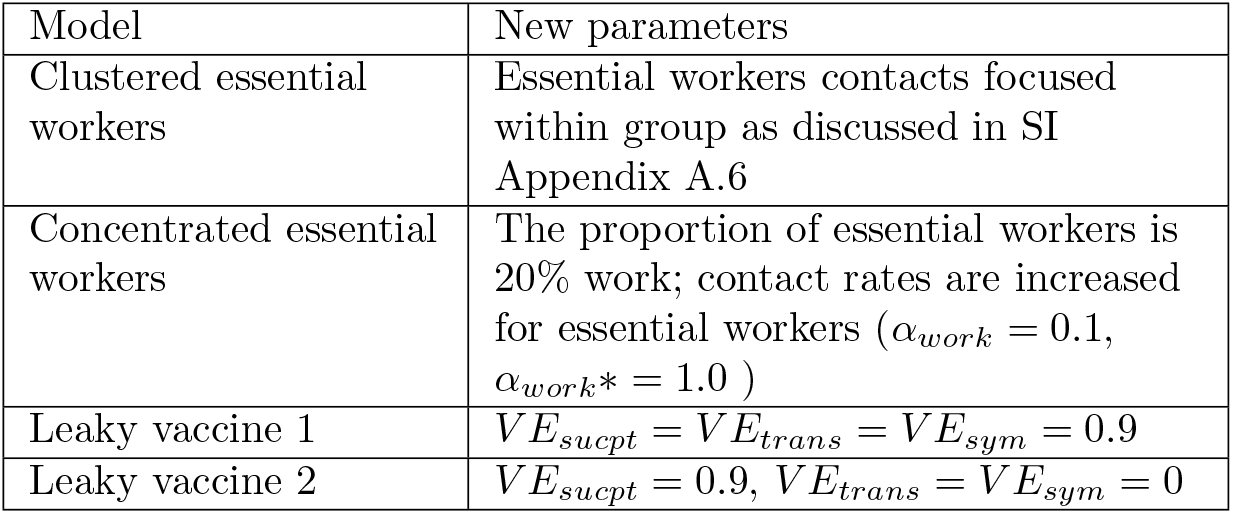
The parameter values changed between the alternative model structures and the base model.

**Figure 10:**
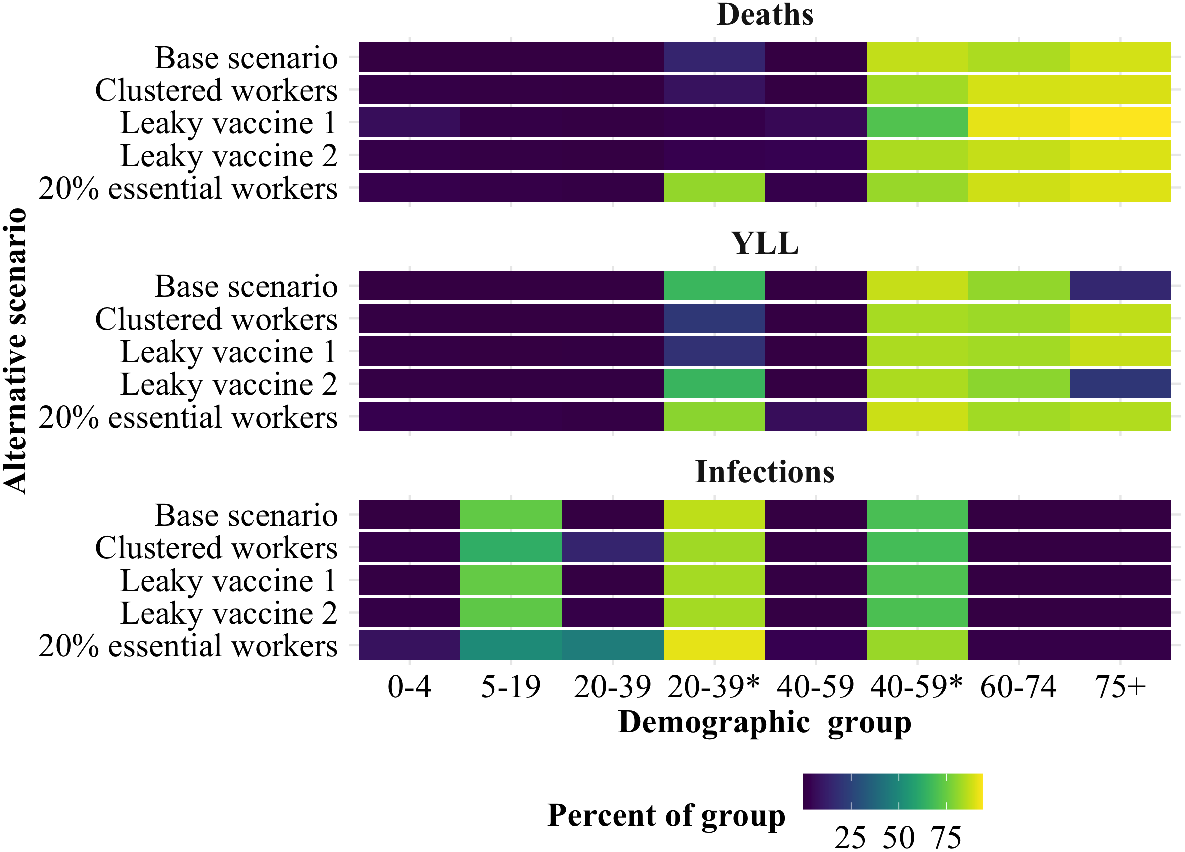
The percent of each demographic group (horizontal axis) vaccinated after three months under the optimal policy for each of the alternative model structures (vertical axis) and objectives (panels).

#### D.1 Leaky vaccine specification

Vaccines can provide multiple forms of protections against infections. Among these protections is the ability for vaccines to prevent individuals from becoming infected (the case considered in the main text). In addition, if vaccinated individuals still become infected they may (1) be exhibit reduced infectiousness and/or (2) develop less severe symptoms. To allow for these latter two cases we changed the model structure to track vaccinated and infected individuals. To do this we maintained the protected and uninfected category *P* and added four categories: vaccinated and exposed class *P*_*exp*_, vaccinated and pre-symptomatic *P*_*presym*_, vaccinated and asymptomatic *P*_*asym*_ and vaccinated and symptomatic *P*_*sym*_. The effectiveness of the vaccine is modeled with three age specific vectors, *V E*_*sucpt*_, *V E*_*trans*_, and *V E*_*sym*_, which quantify the extent to which the vaccine reduces the susceptibility of vaccinated individuals to infection, the reduction in infectiousness of vaccinated individuals and the reduction in infection fatality rate of vaccinated individuals. This new model can be described by the following system of equations:

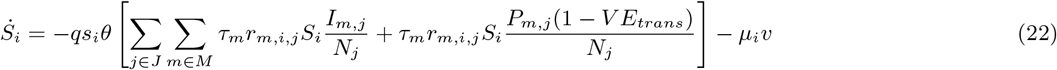

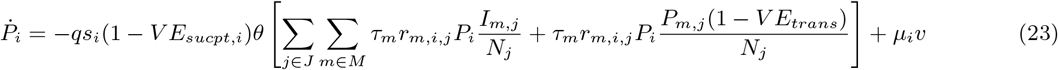

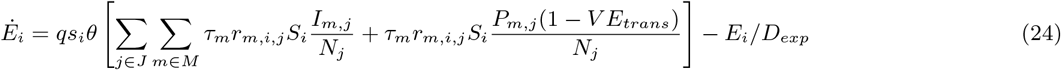

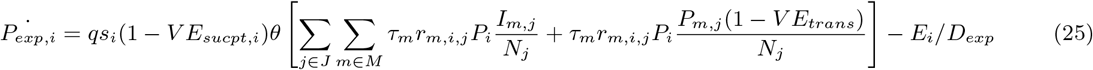

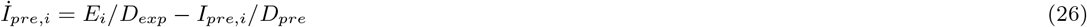

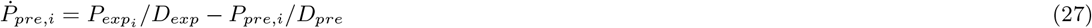

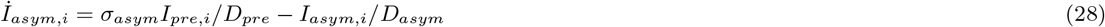

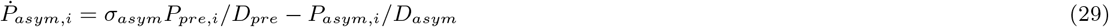

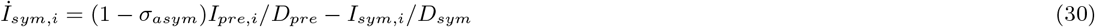

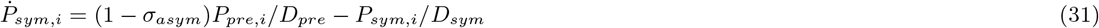

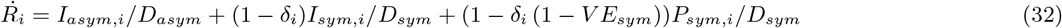

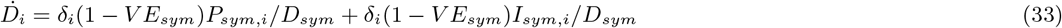

We consider two cases: a vaccine with equal effectiveness set to 90% for consistency with the Base model *V E*_*sym*_ = *V E*_*sucpt*_ = *V E*_*trans*_ = 90%, and a vaccine that only reduces susceptibility to infections *V E*_*sym*_ = 90% and *V E*_*sucpt*_ = *V E*_*trans*_ = 0%.

#### D.2 Contact rates sensitivity

One key source of both uncertainty and heterogeneity between communities is the true set of underlying social contact rates. To test the effects of these parameters we considered a range of work and other contact rates around those specified in the Base scenario. The “other” (outside of the home, school and workplace) contacts were scaled from the pre-COVID-19 average while the work contact rates were increased for essential workers and held at 10% of pre-COVID-19 levels for non-essential workers. Results are presented in Fig. 11, where each scenario is labeled with the percentage change in average work contact rate and the contact rates of essential workers are a percentage of pre-COVID-19 average levels. We found that the policies did change as both other and work contacts increased and that these changes were consistent with the changes in reproductive number induced by the higher contact rates.

**Figure 11:**
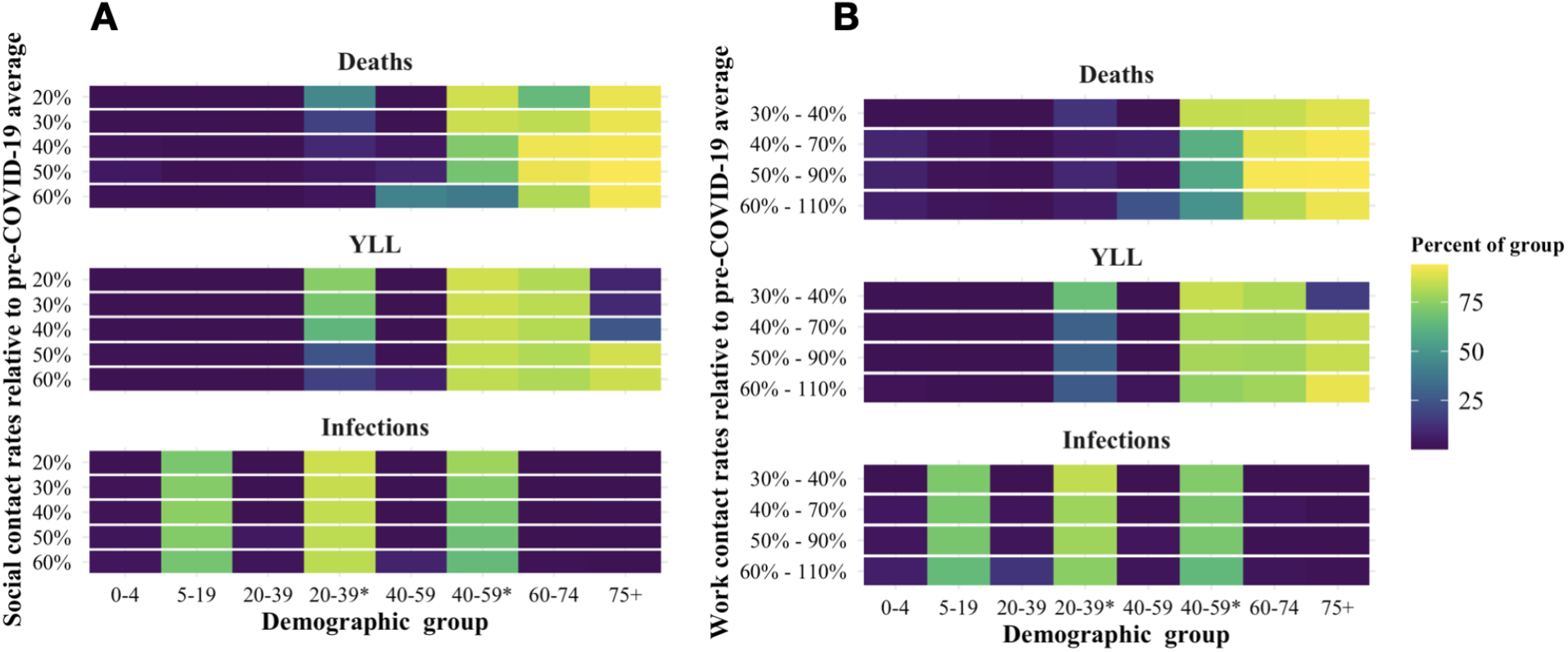
The percentage of each demographic group vaccinated after 3 months under the optimal dynamic policy given variation in (A) “other” (non-work) contacts and (B) work contacts.

## Footnotes

1 We also note (18) use simulation without optimization to explore implications of vaccines with various levels of direct and indirect protection.

2 Excess vaccine is allocated without targeting if all the susceptible individuals in a given group have already been vaccinated.

3 See resulting YLL and resulting infections for the deaths objective with age-only and static constraints.

4 In the event that vaccination requires two doses over time, we consider an individual vaccinated upon receipt of the second dose at time *t* and we assume that *v* indicates the number of *individuals* that can be vaccinated with the required number of doses.

5 This vaccine effectiveness is inclusive of any efficiency loss from typical handling in the distribution chain.

6 If all of the susceptibles from a single group were exhausted (either by full coverage from the vaccine or from infection) then vaccine that would have been allocated to individuals from that group are instead allocated to other age groups at a rate proportional to the size of their susceptible population.

7 It should be noted that the quantities plotted in Fig. 7A correspond to the allocation of the initial supply, which is different from the similar main text Fig. reffig:heatmap_*r*_*obustnessthatpresentsthepercentofeachgroupvaccinatedat*3*months*.

